# Artificial Intelligence in Periodontology: Performance Evaluation of ChatGPT, Claude, and Gemini on the In-service Examination

**DOI:** 10.1101/2024.05.29.24308155

**Authors:** Bushra Ahmad, Khaled Saleh, Saleh Alharbi, Hend Alqaderi, Y. Natalie Jeong

## Abstract

**Background:** Artificial intelligence (AI) language models have shown potential as educational tools in healthcare, but their accuracy and reliability in periodontology education require further evaluation. In this study we aimed to assess and compare the performance of three prominent AI language models—ChatGPT-4o, Claude 3 Opus, and Gemini Advanced—with second-year periodontics residents across the United States on the American Academy of Periodontology 2024 in-service examination.

**Methods:** We conducted a cross-sectional study using 331 multiple-choice questions from the 2024 periodontology in-service examination. We evaluated and compared the performances of ChatGPT-4o, Claude 3 Opus, and Gemini Advanced across various question domains. The results of second-year periodontics residents served as a benchmark.

**Results:** ChatGPT-4o, Gemini Advanced, and Claude 3 Opus significantly outperformed second-year periodontics residents across the United States, with accuracy rates of 92.7 percent, 81.6 percent, and 78.5 percent, respectively, compared to the residents’ 61.9 percent. The differences in performance among the AI models were statistically significant (*p* < 0.001). Percentile rankings underscored the superior performance of the AI models, with ChatGPT-4o, Gemini Advanced, and Claude 3 Opus placing in the 99.95th, 98th, and 95th percentiles, respectively.

**Conclusion:** ChatGPT-4o displayed superior performance compared to Claude 3 Opus and Gemini Advanced. The results highlight the potential of AI large language models (LLMs) as educational tools in periodontology and emphasize the need for ongoing evaluation and validation as these technologies evolve. Researchers should explore both the integration of AI language models into periodontal education and their impact on learning outcomes and clinical decision-making.

## 1 INTRODUCTION

Artificial intelligence is revolutionizing health care, including dentistry, with language models such as Chat Generative Pre-Trained Transformer (ChatGPT), that have shown impressive capabilities in natural language processing and generation, making them valuable educational tools for healthcare professionals.[1,2,3] Large language models (LLMs), including ChatGPT, Claude, and Gemini, are trained on extensive text data, enabling them to generate human-like language with proficiency, thus offering potential advancements in healthcare education by providing intelligent assistance and personalized learning experiences.[4,5] However, integrating LLMs into dental education requires careful evaluation to ensure accuracy, reliability, and adherence to evidence-based practices.[6,7]

In the field of periodontology, AI-powered chatbots could potentially serve as valuable resources for residents and practitioners seeking quick access to information and guidance. These models can assist in various tasks, such as answering clinical questions, thereby providing evidence-based recommendations, and offering interactive learning experiences.[8] For example, AI chatbots could help residents quickly retrieve information about periodontal disease classifications, treatment protocols, and medication dosages, saving time and effort in searching through textbooks or online resources.[9] Additionally, these models could facilitate case-based learning by presenting virtual patient scenarios and guiding users through the diagnostic and treatment planning processes.[10]

However, concerns have been raised regarding the accuracy and reliability of the information provided by these models.[11] Because AI language models are trained on vast amounts of data from the internet and other sources, they may be susceptible to biases, inconsistencies, and misinformation.[12] This is particularly concerning in the context of healthcare education, where the dissemination of inaccurate or misleading information could have serious consequences for patient care.[13] Therefore, it is crucial to evaluate AI language models’ performance and limitations rigorously before integrating them into educational programs or clinical decision support systems.

Previous researchers have examined the performance of ChatGPT, one of the most prominent AI language models, on various healthcare-related assessments, revealing varying levels of proficiency across different disciplines. In the field of neurosurgery, ChatGPT 4 demonstrated impressive performance on written board examinations, achieving a score of 83.4 percent.[14] Similarly, Mihalache et al. evaluated ChatGPT-4 on ophthalmology board certification practice questions, where it correctly answered 105 out of 125 questions (84 percent).[15] However, researchers have not fully explored the models’ performance in the context of dental education. Danesh et al. recently evaluated ChatGPT’s performance on board-style dental knowledge assessment, finding that although ChatGPT 4 showed improved proficiency compared to its earlier version, both models had limitations in their dental knowledge.[16]

To date, no study has comprehensively evaluated the performance of the three largest commercially available AI models on periodontology-specific assessments. Understanding the strengths and limitations of these models in the context of periodontal knowledge is crucial for determining their potential role in education and clinical decision support. Moreover, comparing the performance of different AI models can provide insights into their relative capabilities and inform the selection of the most appropriate model for specific applications.

Therefore, our primary objective in this study is to assess and compare the performance of three prominent AI language models—ChatGPT-4o, Claude 3 Opus, and Gemini Advanced—with the performance of second-year periodontics residents across the nation on the 2024 periodontology in-service examination serving as a benchmark. The periodontology in-service examination is a widely used assessment tool for evaluating periodontal residents’ knowledge and skills in the United States.[17] By analyzing the models’ performance across various topics and types of questions, we aim to provide a comprehensive understanding of their current capabilities and limitations in the field of periodontology.

## 2 MATERIALS AND METHODS

### 2.1 Study Design and Data Collection

Through this cross-sectional study, we evaluated the performance of three AI language models— ChatGPT-4o, Claude 3 Opus, and Gemini Advanced—using the performance of second-year periodontics residents as a benchmark on the 2024 periodontology in-service examination administered by the American Academy of Periodontology (AAP). We obtained the examination questions through the AAP’s website. We conducted the study using the latest versions of the AI models available as of May 21, 2024. Institutional review board approval was not required for this study, as it did not involve human subjects and utilized publicly available AI models and examination questions.

### 2.2 Artificial Intelligence Language Models

ChatGPT-4o, developed by OpenAI, is an advanced language model known for its strong performance across various domains.[18] It has demonstrated impressive capabilities in natural language understanding, generation, and reasoning. Claude 3 Opus, created by Anthropic, is another state-of-the-art AI model that has shown remarkable language understanding and generation abilities.[13] Gemini Advanced, developed by Google, is an LLM that has shown promising results in various natural language processing tasks.[19]

### 2.3 Data Analysis

We used 331 multiple-choice questions from the 2024 periodontology in-service examination to evaluate the AI models. The questions covered various topics, including embryology and anatomy/ biostatistics (n = 29), biochemistry and physiology (n = 31), microbiology and immunology (n = 25), periodontal etiology and pathogenesis (n = 37), pharmacology and therapeutics (n = 36), diagnosis (n = 29), treatment planning and prognosis (n = 27), therapy (n = 77), and oral pathology/medicine (n = 40).

We input the multiple-choice questions and their options (i.e., A, B, C, or D) into the AI models exactly as they appeared in the examination. We determined the correct answers based on the answer key the AAP provided.

To investigate the performance of ChatGPT-4o, Claude 3 Opus, and Gemini Advanced on the AAP in-service exam, we presented each question to the AI models, prompting them with the question: “What is the answer?” We limited the models to a single attempt per question. All questions, including those with clinical pictures, histology slides, schematic images, radiographs, and electrocardiogram (ECG), were included in the study. During data collection, we recorded the AI-generated responses to each question and noted whether the question included clinical data.

The results of the periodontics residents’ performance in the AAP in-service examination were obtained from the periodontology department at Tufts School of Dental Medicine.

We performed a descriptive analysis to evaluate the number and percentage of correct answers. The chi-square test was used as a test of association between categorical variables. We used IBM’s SPSS Statistics 29 and GraphPad’s Prism software to analyze and graph the data.

## 3 RESULTS

Table 1 compares the performance of three AI models – ChatGPT-4o, Claude 3 Opus, and Gemini Advanced - across various domains related to AAP in-service exam questions. The table presents the number and percentage of correct answers given by each model in each domain. ChatGPT-4o demonstrates the highest overall performance, with accuracy ranging from 85.7 percent to 100% across all domains. Claude 3 Opus and Gemini Advanced show slightly lower performance compared to ChatGPT-4o, with accuracy ranging from 57.1 percent to 100 percent and 66.2 percent to 100 percent, respectively. ChatGPT-4o outperforms the other two models in most domains, with notable differences in the therapy as well as treatment planning and prognosis domains.

**TABLE 1:** Comparative Performance Analysis of ChatGPT-4o, Claude 3 Opus, and Gemini Advanced AI Models in AAP 2024 In-Service Examination Question Domains.

| Domain | ChatGPT-4o<br>N (%) | Claude 3 Opus<br>N (%) | Gemini<br>Advanced<br>N (%) |
| --- | --- | --- | --- |
| Biochemistry-Physiology | 30/31 (96.8) | 28/31 (90.3) | 30/31 (96.8) |
| Diagnosis | 29/29 (100) | 25/29 (86.2) | 22/29 (75.9) |
| Embryology and<br>Anatomy /<br>Biostatistics | 27/29 (93) | 22/29 (76) | 25/29 (86) |
| Microbiology and<br>Immunology | 25/25 (100) | 23/25 (92) | 23/25 (92) |
| Oral Pathology/<br>Oral Medicine | 37/40 (92.5) | 35/40 (87.5) | 36/40 (90) |
| Periodontal<br>Etiology and<br>Pathogenesis | 36/37 (97.3) | 32/37 (86.5) | 32/37 (86.5) |
| Pharmacology and<br>Therapeutics | 33/36 (91.7) | 33/36 (91.7) | 31/36 (86.1) |
| Therapy | 66/77 (85.7) | 44/77 (57.1) | 51/77 (66.2) |
| Treatment<br>Planning and<br>Prognosis | 24/27 (88.9) | 18/27 (66.7) | 20/27 (74.1) |

Figure 1 presents a visual comparison of the overall performance of three large LLMs— ChatGPT-4o, Gemini Advanced, and Claude 3 Opus—along with the average performance of second-year periodontics residents across the United States. The bar graph clearly illustrates that all three LLMs outperformed the second-year periodontics residents, with ChatGPT-4o achieving the highest accuracy at 92.7 percent, followed by Gemini Advanced at 81.6 percent, and Claude 3 Opus at 78.5 percent, whereas the second-year periodontics residents achieved 61.9%. The performance differences among the LLMs were statistically significant *(p* < 0.001). Furthermore, the percentile rankings demonstrate the superior performance of the AI models, with ChatGPT-4o, Gemini Advanced, and Claude 3 Opus placing in the 99.95th, 98th, and 95th percentiles, respectively, while the second-year residents ranked in the 50th percentile.

**F I G U R E 1:**
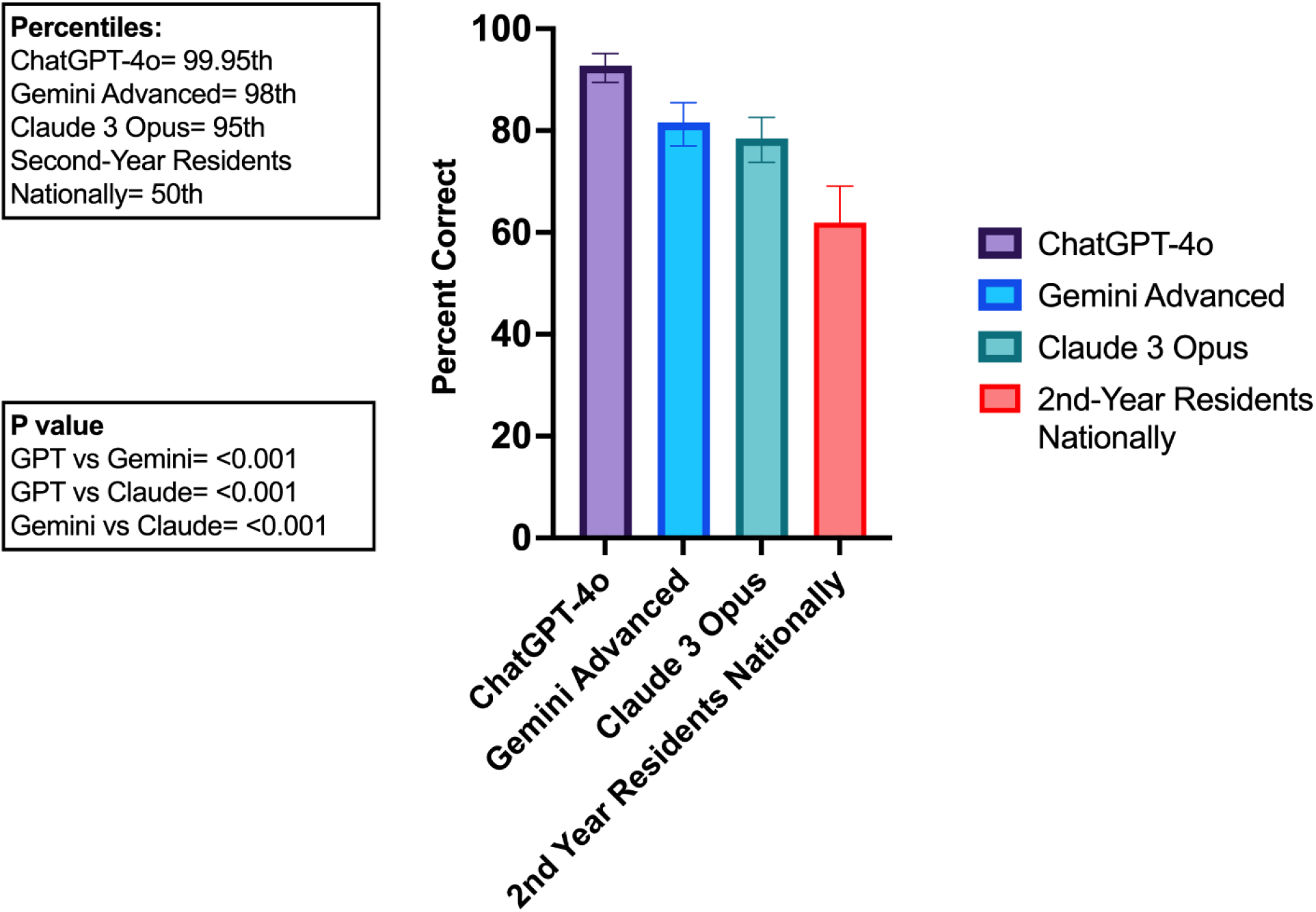
Comparative Performance of Different Large Language Models Against Second-Year Periodontics Residents Across the United States in the AAP 2024 In-Service Examination

Figure 2 illustrates the performance of AI models across different types of questions, including radiographs, histology slides, clinical images, schematic images, and ECGs. All AI models correctly answered the radiograph and ECG questions. For the histology questions, all models answered two out of three correctly. In the case of clinical images, the models also achieved a correct response rate of two out of three correct response rates. Regarding schematic images, both Gemini Advanced and Claude 3 Opus answered 2 out of 3 questions correctly, whereas ChatGPT-4o managed to answer one out of three correctly.

**F I G U R E 2:**
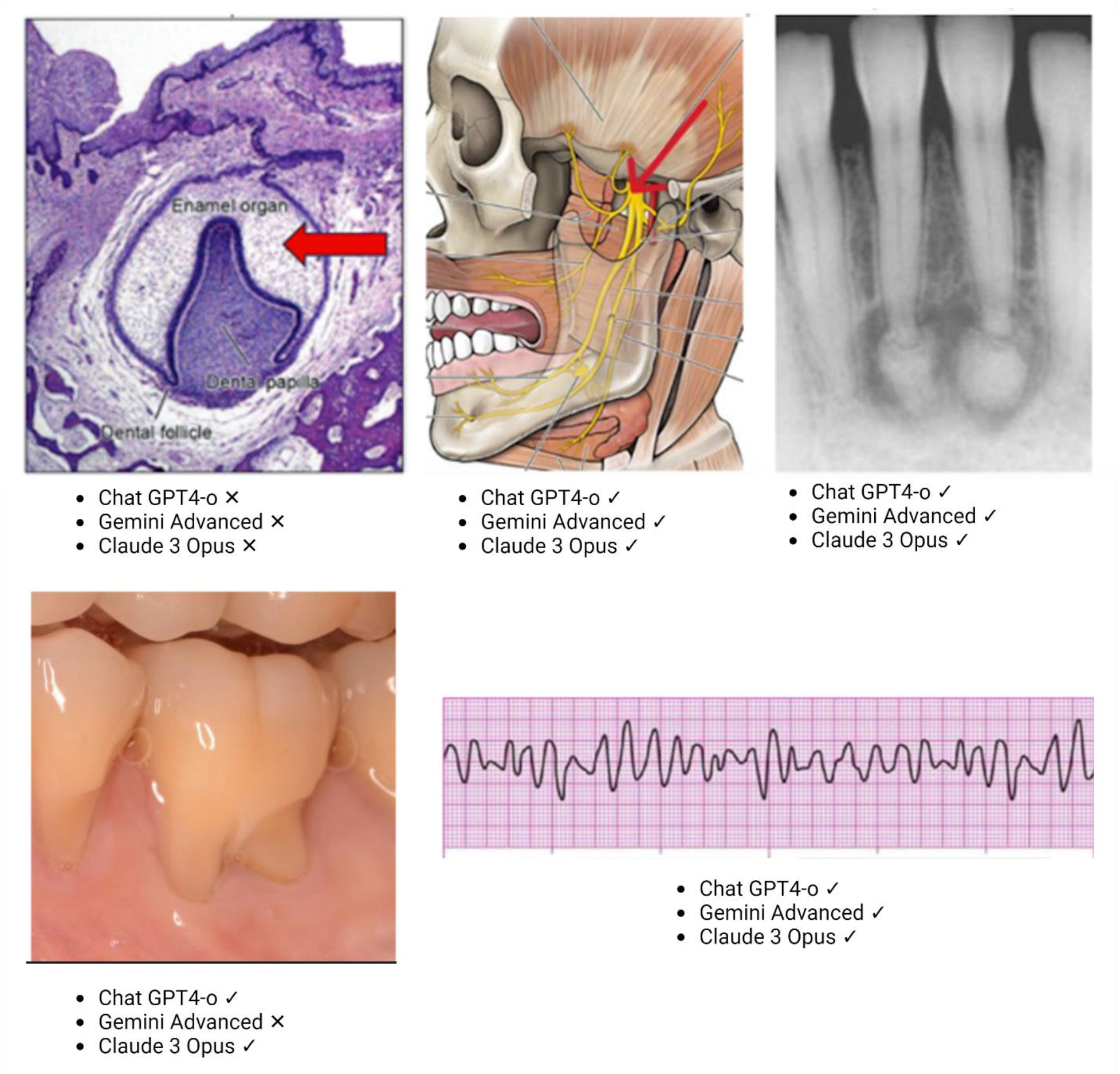
Distribution of question types: radiographs, histology slides, clinical images, schematic images, and electrocardiograms.

## 4 DISCUSSION

Through this study, we evaluated the performances of ChatGPT-4o, Claude 3 Opus, and Gemini Advanced on the AAP 2024 in-service examination, demonstrating these AI models’ superior performances compared to second-year periodontics residents across the United States. ChatGPT-4o achieved the highest accuracy across all domains, ranging from 85.7 percent to 100 percent, significantly outperforming not only Claude 3 Opus and Gemini Advanced but also the periodontics residents.

Including the performances of second-year periodontics residents as a benchmark highlights the potential of advanced AI language models to support and enhance periodontics education because the models consistently outperformed the residents. However, it is important to recognize that AI models are not replacements for human expertise and clinical judgment because they may lack the nuanced understanding and contextual awareness that experienced clinicians possess. The results should be interpreted as a testament to AI’s potential to augment periodontics education rather than as a direct comparison of machine and human capabilities.

Our study, which evaluated the performance of ChatGPT-4o, Claude 3 Opus, and Gemini Advanced on the AAP 2024 in-service examination, shows that ChatGPT-4o achieved the highest accuracy across all domains, ranging from 85.7 percent to 100 percent, significantly outperforming Claude 3 Opus and Gemini Advanced, which had lower accuracy rates in several domains. In contrast, in their study on the performances of ChatGPT-3 and ChatGPT-4 on the 2023 AAP in-service examination, Danesh et al. found that ChatGPT-4 answered 73.6 percent of the questions correctly, whereas ChatGPT-3.5 managed only 57.9 percent.^**20**^ This indicates that ChatGPT-4o demonstrated a significant improvement in performance, outperforming both ChatGPT-4 and ChatGPT-3.5 with higher accuracy rates across all domains.

In our study, we included all 331 questions in the analysis, with no exclusions. By incorporating questions with clinical pictures, histology slides, schematic images, radiographs, and ECGs, we provided a comprehensive assessment of the AI models’ capabilities. We utilized the three largest commercially available AI models, all of which can interpret images, marking a significant strength of our study. This approach contrasts with that of Danesh et al., who excluded image-based questions owing to ChatGPT 3.5 and 4 models’ limitations in processing visual inputs.[20] Our findings indicate that although all three models excelled in interpreting the radiograph and ECG, their performance varied for histology slides, clinical images, and schematic images. This suggests that the models’ abilities to process and analyze visual information may be an area for further improvement and research.

Our study also highlights the ChatGPT-4o’s superior performance in the therapy and treatment planning and prognosis domains compared to the other models. This is a significant finding because these domains are crucial for clinical decision-making and patient care. ChatGPT-4o’s ability to provide accurate and reliable information in these areas could potentially support dental professionals in making evidence-based treatment decisions. However, it is important to emphasize that AI language models are not substitutes for clinical judgment and that trained professionals should critically evaluate their outputs.[21]

It is worth noting that AI language models’ performance is continually evolving through updating and fine-tuning on new data. Therefore, the results of our study should be interpreted in the context of the specific model versions used (ChatGPT-4o, Claude 3 Opus, and Gemini Advanced) and the time at which we conducted our study. As these models continue to advance, it will be important to reassess their performance and capabilities regularly.

One potential limitation of our study is the use of a single assessment tool, the AAP 2024 in-service examination, to evaluate the AI models’ performances. Although this examination is widely used and respected in the field of periodontology, it may not fully capture the breadth and depth of knowledge required for clinical practice. Future studies could consider using a range of assessment methods, such as case-based scenarios or clinical simulations, to provide a more comprehensive evaluation of the models’ capabilities.[22]

## 5 CONCLUSION

Our study revealed that ChatGPT-4o demonstrated superior performance, with accuracy rates ranging from 85.7 percent to 100 percent, significantly outperforming Claude 3 Opus and Gemini Advanced. Compared to second-year periodontics residents across the United States, all three AI models showed superior performance, underscoring their potential to support periodontal education and clinical decision-making.

However, we emphasize the need for ongoing evaluation and validation of AI models across diverse periodontal topics and question types, as well as the importance of trained professionals critically evaluating their outputs. The findings have implications for the future development and integration of AI in periodontal education and practice, underlining the necessity of collaboration between AI developers, dental educators, and clinicians to ensure the responsible and effective use of these technologies. As AI continues to advance, fostering open communication and collaboration among stakeholders is crucial to maximize the benefits of AI in periodontal education and ultimately improve patient outcomes.

## Author Approval

All authors have seen and approved the manuscript.

## Data Availability Statement

The data that supports the findings of this study are available from the corresponding author upon reasonable request.

## AUTHOR CONTRIBUTIONS

Bushra Ahmad designed the study, acquired, and interpreted the data, helped with data analysis, and wrote the manuscript. Khaled Saleh acquired and interpreted the data and wrote the manuscript. Saleh Alharbi acquired the data and revised the manuscript. Hend Alqaderi helped with revising the manuscript. Y. Natalie Jeong was involved in gathering the data and revising the manuscript critically. All authors approved the final version of the manuscript to be published and agreed to be accountable for all aspects of the work.

